# Study protocol for Running for Health (Run4Health): a randomized controlled trial investigating the effect of Frame Running (RaceRunning) training on cardiovascular health in children and youth with cerebral palsy

**DOI:** 10.1101/2023.08.06.23293736

**Authors:** Sarah E Reedman, Ellen L Armstrong, Leanne Sakzewski, Stina Oftedal, Matthew Ahmadi, Andrea Burgess, Tamara Blake, Syed Afroz Keramat, Iain Dutia, Dayna Pool, Lynda McNamara, Rachel Thomas, Kerry West, Stewart G Trost, Mark Peterson, Emma Beckman, Catherine Sherrington, Tracy Comans, Robert Ware, Fiona Russo, Craig Munns, Kristie Bell, Laura Gascoigne-Pees, Denise Brookes, Roslyn N Boyd

## Abstract

**Introduction:** It is well established that young people with moderate-severe (Gross Motor Function Classification System [GMFCS] levels II-V) cerebral palsy (CP) participate in less physical activity compared to typically developed peers, and children with CP who can walk without limitations (GMFCS level I). Frame Running (formerly RaceRunning) is a World Para Athletics sanctioned sport that allows people with moderate-severe CP to access, experience and compete in running using a specialised three-wheeled frame with low rolling resistance. The Run4Health pilot randomised controlled trial (protocol published elsewhere) was designed to investigate the cardiorespiratory benefits of a 12-week frame running training program in young people with CP (aged 8-21 years, GMFCS II-V). Following enrolment of 12 participants in the pilot study, additional funding was secured to expand the Run4Health study to include additional training/study sites, new research questions and outcome measures, based on feedback from consumers. Such changes necessitate an expanded and updated study protocol. This expanded Run4Health study will investigate the effects of a 12- week Frame Running training program on cardiorespiratory health, bone mineral density, gross motor function and capacity, physical activity participation, sleep, pain and quality of life in children and youth (aged 8-21 years) with moderate-severe CP (GMFCS levels II-V).

**Methods and Analysis:** One hundred and two children and youth with CP (age 8-21 years) classified in GMFCS levels II-V will be recruited across three sites (six training locations) and randomised to receive either 12 weeks of Frame Running training twice weekly for 60 minutes, or 12 weeks of usual care (waitlist control group). Outcomes will be measured at baseline, immediately post-intervention, and 12 weeks post-intervention. The control group will receive the intervention following T3, and have an additional assessment session following 12 weeks of training (T4). Outcomes include cardiorespiratory fitness, bone mineral density, blood pressure, habitual physical activity, body mass index, waist circumference, percentage body fat, gross motor function and capacity, community participation, sleep, pain, quality of life and mood, health utility, feasibility, tolerability, and safety. Adverse events will be monitored, and participants, caregivers and coaches will be interviewed to explore barriers and facilitators to ongoing, sustainable participation in Frame Running.

**Ethics and Dissemination:** Ethical approval for this study was granted by The Children’s Health Queensland Hospital and Health Service (HREC/21/QCHQ/69281) and the University of Queensland Human Research Ethics Committees (2021/HE000725). Research outcomes will be disseminated via scientific conferences and publications in peer reviewed journals; to therapists and coaches through professional and athletic organisations; and to people with CP and their families.

**Registration:** Australian New Zealand Clinical Trials Registry number: ACTRN12621000317897

## INTRODUCTION

Frame Running (RaceRunning) is a Para athletics discipline that provides an accessible physical activity option for people with moderate-severe cerebral palsy (CP). A specialized running frame with low rolling resistance allows athletes with severe mobility limitations to assume a supported running position and to self-propel on a sealed running track. Although findings from an adequately powered randomized controlled trial (RCT) are not yet available, results from a pre-post pilot study including 15 young people with CP suggest that Frame Running training twice per week for 12 weeks, led to improvements in cardiorespiratory endurance and gastrocnemius muscle thickness^1^. People with CP who participated in Frame Running have also demonstrated heart rates commensurate with high intensity exercise, providing further support for Frame Running as a means to improve cardiovascular fitness in this population.^2^

Run4Health CP is an ongoing, adequately powered, RCT investigating the effects of Frame Running training on cardiometabolic risk factors in young people with moderate-severe CP.^3^ The recently published Run4Health CP protocol includes a detailed 12-week training program, featuring two 60-minute training sessions per week conducted at community-based athletics tracks.^3^ As of July 2023, twelve children and adolescents with CP, classified in levels II-V on the Gross Motor Function Classification Scale [GMFCS] had enrolled and participated in the pilot study. Additional research funding (secured after the enrolment of 12 pilot participants), has allowed the Run4Health RCT to be expanded to include: a larger study cohort (N=102, including participants enrolled in the pilot study), an additional research site, three new training locations, the inclusion of a waitlist control group and the addition of measures of lung function, sleep, bone mineral density (BMD) and a selection of participant self-report outcomes. The new research questions and outcomes were included following consultation with the Run4Health consumer group. Although the training program will remain unchanged, expansion of the Run4Health study to include additional research questions, outcome measures, training sites and assessment timepoints, necessitates this updated study protocol.

This updated Run4Health study will investigate whether the Run4Health training program leads to improvements in lung function, bone mineral density and body composition, physical activity participation, community participation, functional strength, mental health, quality of life, sleep and gross motor function in young people with CP (aged 8-21 years), GMFCS II-V.

### Justification to include additional outcome measures in the expanded Run4Health RCT

Young people with CP experience reduced cardiorespiratory fitness compared to typically developing peers, which is associated with low levels of physical activity participation and high levels of sedentary behaviour.^4^ This is concerning because respiratory and cardiovascular diseases and illnesses are among the leading causes of death in young people with CP.^5 6^ Adults with CP also have a much higher likelihood of developing a non-communicable disease associated with low PA compared to the typically developed population.^4^

Increased physical activity likely increases vital capacity, optimises chest wall mobility, and promotes small airway clearance in people with CP.^6^ A systematic review and consensus statement on the prevention and management of respiratory disease in people with CP identified 3 uncontrolled studies of exercise interventions (a breathing exercise program^7^, upper limb resistance training with elastic bands^8^ and an aquatic and gym exercise program^9^) that led to improved lung function in ambulant children with CP.^6^ These interventions, however, had limited accessibility to youth with moderate-severe CP (GMFCS II-V), due to poor adherence, limited upper limb function, and contraindications to aquatic exercise.^6^

Measuring respiratory muscle strength before and after the Run4Health training program, using feasible and accurate techniques, would provide important information on lung health in children with moderate-severe CP (GMFCS II-V).

Traditional lung function measures (i.e., spirometry, lung volumes) require high levels of effort to produce acceptable results.^10^ Children with CP have loss of respiratory muscle function resulting in an ineffective cough, decreased ventilation and reduced lung compliance.^11^ Techniques to measure diaphragm (maximal inspiratory pressures [MIPS], sniff nasal inspiratory pressures [SNIPs]) and abdominal/intercostal muscles (maximal expiratory pressures [MEPS]) strength have been shown to be feasible in people with neuromuscular diseases.^12^ New techniques, such as intra-breath oscillometry (IB-OSC) are also feasible, as data can be collected through 20-second periods of tidal breathing. IB-OSC measures respiratory system resistance (airway obstruction) reactance (lung compliance), and ventilation homogeneity (evenness of gas mixing).^13^

Further to exhibiting poor cardiorespiratory fitness, children and youth with moderate-severe CP (GMFCS II-IV) have low BMD, altered body composition and increased risk of low trauma fractures. Impaired motor function and poor nutritional status evidenced by low lean mass are significant contributors to the greater risk of low BMD in the CP population.^14^ Low BMD has resulted in a fracture rate of 7-9.7% per year, with the majority being non-traumatic fractures, most frequently in the distal femur.^15^ Prospective CP-child cohort studies have examined neurodevelopment, growth, nutrition, and physical activity in 245 children with CP from 2-5 years to 8-12 years.^16–19^ This longitudinal study found that children classified in GMFCS II-V had poor growth and altered body composition evidenced by shorter stature, lower body weight, lower fat free mass and higher fat mass relative to population-level references, and that sedentary behaviour peaked and plateaued as early as 3 years of age.^16 20^

Frame Running has the potential to promote increased BMD in the lower limbs and spine. There is emerging evidence that weight-bearing exercise interventions in children with CP significantly improves BMD in the femur.^21^ A pre-post pilot study (N=15 children with CP, age 4-12 years, GMFCS IV-V), of Frame Running training for 12 weeks (60 minutes at 3 times weekly) demonstrated improvements in calcaneal bone quality index on quantitative ultrasound.^22^ Trials using reliable measures of BMD such as dual energy x-ray absorptiometry (DXA), are required to more accurately capture potential impacts of Frame Running on body composition.

Children with CP have insufficient and disrupted sleep.^23^ Sleep is a key determinant of health and wellbeing across the lifespan. Increasing physical activity improves sleep outcomes in adults without CP, however there is limited research into the effects of physical activity and exercise interventions on sleep in youth with CP. As such, sleep quality, sleep duration, sleep efficiency, wake after sleep onset, and sleep latency captured using a combination of questionnaire and device-based measures are also included in this expanded Run4Health RCT protocol.

Children with moderate-severe CP have limited ability to increase gross motor capacity and mobility performance over time. A longitudinal study of Australian children with CP found that children classified in GMFCS level V showed no changes in gross motor capacity or mobility performance between 18 months and 12 years of age, while children GMFCS level IV plateaued in capacity and performance by 5 years of age.^25^ A systematic review of adults with CP suggested that 25% of ambulant adults experience a decline in mobility, and greater disability was associated with a higher risk of declining mobility over time.^26^ Those who engaged in regular physical activity were at a lower risk of mobility decline, and gait deterioration was strongly associated with higher sedentary behaviour.^26^ To better capture improvements in gross motor function, mobility and frame running-specific outcomes across the spectrum of GMFCS levels, the updated Run4Health protocol will therefore include: the item set version of the Gross Motor Function Measure (GMFM)^27^; the mobility domain of the Pediatric Evaluation of Disability Index (PEDI-CAT)^28^; and frame running participation and goal attainment, as measured by the Canadian Occupational Performance Measure (COPM).

The decision to update the Run4Health protocol to include these additional outcomes was made in consultation with the Run4Health consumer group and following the successful procurement of an Australian Government Medical Research Future Fund (MRFF) grant (2022624).

## METHODS AND ANALYSIS

### Objectives

The primary study objective is to compare the effect of 12-weeks of Frame Running training, twice weekly, compared to usual care (waitlist control group) on cardiovascular fitness, assessed using i) the Six Minute Frame Runner Test (6MFRT, H1) and ii) 1-minute heart rate recovery (HRR1min) following exercise testing immediately at post-intervention (primary endpoint, T2) and at 12 weeks post-intervention (T3).^3^

Secondary objectives are to compare the effect of 12 weeks of Frame Running training versus usual care (waitlist control group) at T2 and T3 on: iii) gross motor function (GMFM), iv) functional mobility (PEDI-CAT), v) Frame Running specific activity limitation tests (distance covered in four strides, ground contacts in 20 metres and 100m sprint time), vi) resting blood pressure, vii) waist circumference viii) body composition including fat mass, lean body mass, BMD and bone mineral content of the whole body, lateral distal femur and lumbar spine, ix) physical activity, sedentary behavior and sleep duration using device-based measures, x) lung function (inspiratory/expiratory muscle strength, ventilation distribution), xi) sleep quality, xii) self-reported and/or parent-proxy reported mood and quality of life (KIDSCREEN-52), xiii) pain, xiv) cost effectiveness including costs and consequences (Health Resource Use Questionnaire [HRUQ], Medicare Benefits Schedule [MBS] and Pharmaceutical Benefits Scheme [PBS] data) and Health Utility (as assessed by the CHU-9D for children 0-17 years and EQ-5D-5L for youth 18-21 years), and xv) performance and satisfaction for Frame Running activity and participation goals (COPM).

The tertiary aim of this study is to evaluate effectiveness of implementation by identifying the number of athletes who continue to train in the community at 3-month post follow-up, including participation frequency of attendance and level of involvement, and barriers and facilitators to community Frame Running on the Theoretical Domains Framework (TDF) based questionnaire and qualitative interview at study exit.^29^

### Trial Design

Run4Health CP is a pragmatic, single (assessor)-blind randomized waitlist controlled, multi-centre trial with two parallel groups. The primary timepoint is immediately post-intervention (T2, 12 weeks post-baseline) and the secondary timepoint is 12 weeks post-intervention (T3, 24 weeks post-baseline). Participants in the intervention group will exit the study after T3, and the waitlist group will be offered the intervention. The final assessment point for the waitlist control group will be immediately post-training (T4, 36 weeks post-baseline assessments) (Fig. 3).

The Run4Health RCT^3^ will be expanded to include a total of 102 participants across six Australian training sites: Brisbane (n=25), Gold Coast (n= 15), Sunshine Coast (n=17), Cairns (n=15), Perth (n=15) and Sydney (n=15). Frame Running training and related assessments will be conducted at synthetic athletics tracks in the community or nearby associated indoor sports facilities at a time convenient to participants.^3^ Randomisation will be stratified according to GMFCS (II-III/IV-V) and site (Brisbane vs Gold Coast, vs Sunshine Coast vs Cairns vs Perth vs Sydney), with 1:1 assignment to the intervention group (Frame Running training) or waitlist control group (usual care). Recruitment and implementation of this updated protocol will commence in July, 2023 following enrolment of 12 participants to the pilot study.

### Eligibility Criteria

Eligibility and exclusion criteria are as per the Run4Health pilot RCT protocol.^3^ That is, eligible participants must: i) have a diagnosis of cerebral palsy and classified in GMFCS levels II-V; ii) be aged between 8.00 to 21.99 years of age; iii) live within 150km of one of the trial sites; iv) have not engaged in formal Frame Running training within the last 6 months; v) can understand and follow the directions of the coach and assessors for the purposes of training safely and completing outcome measurement in the opinion of the Principal Investigator.^3^

Participants will be excluded if: i) they have had orthopaedic and/or neurological surgery within 6 months prior to baseline or during the study period requiring a period of recovery that would exclude the participant from training for more than one week; ii) have uncontrolled epilepsy, medical fragility, and/or serious precautions not able to be accommodated (e.g. significant history of atraumatic lower limb fractures or sacral pressure injuries etc.) precluding participation in moderate-vigorous intensity Frame Running; and/or iii) if caregiver English language skills are not sufficient to understand the study information, provide informed consent and/or complete study questionnaires.^3^

### Interventions

#### Frame Running Training (Intervention Group)

The detailed Frame Running training protocol, including the rationale, training intensity, dose and progressions is included in the Run4Health pilot study protocol.^3^ Participants allocated to the training group will complete two 60 minute sessions per week for 12 weeks (total dose of 24 hours), which is considered an adequate dose to improve aerobic fitness in deconditioned individuals with CP.^1, 30^ Training sessions will consist of a combination of anaerobic and aerobic Frame Running, and task-specific functional training for Frame Running technique and skills. The target heart rate for training sessions will be between 116- 185 beats per minute (based on an estimated peak heart rate of 194 beats per minute), which will be monitored using a Polar Verity Sense (Polar Electro Oy, Kempele Finland) optical heart rate monitor on the non-dominant upper limb.^31^ Participants will train in small groups (approximately 2-4), and sessions will be administered by a coach with qualifications in Physiotherapy and/or Exercise Physiology. Participants in the training group may continue to receive any usual care throughout the trial, without restriction.^3^

#### Usual Care (Waitlist Control Group)

Participants in the control group will continue any usual care (as per concomitant care), however they will be asked to refrain from participating in Frame Running until the waitlist period is complete (at T3). The waitlist control group will be offered the 12-week training program following T3, and final assessments will be completed immediately post training, at 36 weeks (T4). It is expected that few participants in the control group will participate in regular Frame Running during the control period, as this would require access to their own running frame, which are not readily available in the community.

#### Modifications and Adaptations

Session difficulty will be monitored and progressed in real-time and from session-to-session based on participants heart rate responses and participant/family feedback following training sessions (e.g. training related soreness). Any excessive or undue pain or fatigue will be reported as an adverse event and may necessitate modifications to the training dose, delivery or content. Modifications and adaptations to the training program will be made as outlined in the Run4Health pilot RCT protocol.^1^ Training sessions will be tailored to facilitate participation of children across various age groups, motor types/distributions, interests, as well as participants with co-diagnoses including (but not limited to) visual impairments, intellectual disability and proprioceptive impairments.

#### Adherence and Fidelity

The Run4Health Training manual, included in the Run4Health pilot RCT protocol^3^, will continue to be implemented by coaches across trial sites to promote adherence to the prescribed training dose. The Run4Health Assessment manual, updated to include the new outcome measures, will be provided to blinded assessors to encourage consistent assessment procedures across trial sites.

The following strategies, underpinned by Self Determination Theory^32^ and explained in greater detail in the pilot RCT protocol^3^, will be implemented to optimize participant’s attendance and involvement in the training program:

i. Training sessions will be tailored to the individual to facilitate a ‘just right challenge’ (i.e. at a level of difficulty that is not too easy and not too difficult).
ii. Participants will train in small groups (preferably with similar ages/abilities) to encourage social connection and to help meet participants’ need for relatedness.^33^
iii. Coaches will endeavour to create an environment that fosters participants’ self-efficacy, by facilitating positive peer interactions, modelling positive self-talk and communicating with participants and families in a way that encourages autonomy.^34^

To monitor adherence to the training program from session to session, coaches will record the percentage of: i) sessions (or part sessions) attended, ii) training drills completed as per the training manual, and iii) session duration spent within the training target-heartrate threshold. Coaches will also record any modifications or adaptations made to the session content (and reason/s for the changes). Missed or incomplete sessions will be recorded and reported alongside study outcomes.^3^

#### Concomitant Care

As per procedures outlined in the Run4Health pilot protocol^3^, participants in both groups may continue any usual care throughout the study period except Frame Running in the waitlist control group during the waitlist period (T1-T3). A health resource utilisation questionnaire will be completed to record frequency of participation in any therapies and physical activities (including Frame Running) throughout the study.

### Outcomes

Study outcomes, except for DXA scans, will be measured at baseline (T1, 0 weeks), immediately post-intervention (T2, 12 weeks, primary comparison) at 12 weeks follow-up (T3, 24 weeks, retention of effects) and at T4 (36 weeks, waitlist group only). DXA scans will be completed at T1 and T2 only, to minimise exposure of participants to radiation. The primary study comparison will be at T2, however outcomes will also be pooled for both groups to assess pre-post intervention change which may allow for increased power to detect difference on the secondary outcomes. This will be useful for exploratory analysis and to understand factors affecting participant’s responses to the intervention.

#### Primary Outcome

The primary study outcome is distance (metres) covered in the Six Minute Frame Runner Test (6MFRT).^35^ The 6MFRT is a valid measure of Frame Running endurance with good test-retest reliability (ICC=0.78-0.91) in children classified in GMFCS levels III and IV.

The following secondary outcomes will be implemented as per procedures outlined in the Run4Health pilot study protocol^3^:

i. Heart rate recovery in 1 minute (HRR_1min_)^36^
ii. Resting blood pressure (mmHg)
iii. Habitual physical activity quantified using accelerometry^37^
iv. Body composition: body mass index (BMI, kg/m^2^) and waist circumference (cm)
v. Gross motor function assessed using the Gross Motor Function Measure (GMFM- 66)^27^
vi. Frame Running functional strength assessments (activity-limitation tests), assessed using a modified functional test battery for children with CP^38^ (100m sprint time, step count in 20 metres and distance in 4 strides)
vii. Community participation evaluated using the Participation and Environment Measure for Children and Youth (PEM-CY)^39^
viii. Feasibility, tolerability and safety as measured by the Wong-Baker FACES® rating scale (pain)^40^, Fatigue Severity Scale (FSS, fatigue),^41^ and training load (Rate of Perceived Exertion [RPE] on the OMNI RPE^42^ multiplied by session duration).^43^

In addition to the above listed outcomes, the expanded Run4Health RCT will also include the following new secondary outcomes:

i. Body composition: Dual energy x-ray absorptiometry (DXA) is a three-compartment measure of bone and body composition that derives bone mineral content and soft tissue mass separately and estimates the latter into fat and lean body mass. Age/height matched areal bone mineral density (aBMD; g/cm2), bone mineral content (BMC; grams) at total body, AP lumbar spine, and (+/-) lateral distal femurs (LDF)^44^, lean and fat mass are reproducible in people with CP. Both lateral distal femora are scanned and aBMD subregions averaged. Lateral distal femur scans may not be measured at all facilities, as this scan site is not a standard clinically reported outcome. Lumbar spine is used to calculate bone mineral apparent density (BMAD, g/cm3), from the projected bone area (cm2) to provide volumetric BMD.^44^ Estimated time for DXA scans is 25 minutes at T1-T2 only. The total radiation dose for the DXA scans (two in total) is about 0.1 mSv^1^. As part of everyday living, everyone is exposed to naturally occurring background radiation and receives a dose of about 2 millisieverts (mSv) each year. At this dose level, no harmful effects of radiation have been demonstrated as any effect is too small to measure. Female participants who are 12 years and over will be asked immediately prior to the scan if there is any chance they could be pregnant, however scanning is not contraindicated and pregnancy testing is not required due to the low dose of radiation. Bone and soft tissue phantoms will be used to standardise between sites. DXA scans will be used in place of callipers to assess percentage body fat.^48^
ii. Sleep: Sleep duration, efficiency, wake after sleep onset and sleep latency will be analysed from wrist accelerometer data using algorithms validated in children with CP.^49^ Pittsburgh Sleep Questionnaire Index (PSQI) is a self-rated questionnaire which assesses sleep quality and disturbances (quality, sleep latency, sleep duration, habitual sleep efficiency, sleep disturbances, use of sleeping medications and daytime dysfunction) over a 1-month period. A global PSQI score >5 yielded a diagnostic Sensitivity 89.6% and Specificity 86.5% (kappa = 0.75, p<0.001) in distinguishing good and poor sleepers (healthy vs. sleep disorder).^50^ To capture physical activity, sedentary and sleep data, wrist and thigh accelerometers will be worn at all times except when the participant is submerged in water (e.g. swimming). Participants will record sleep and wake times and any times that the activity monitors were removed (e.g. for showering) in an activity monitor data logbook.
iii. Lung function: Maximal inspiratory pressures (MIPS), sniff nasal inspiratory pressures (SNIPs) and abdominal/intercostal muscles (maximal expiratory pressures [MEPS] will be performed using the Carefusion MicroRPM device according to American Thoracic Society/European Respiratory Society standards. Participants will perform a maximum inhalation or exhalation manoeuvres against a known resistance. A minimum of 3 acceptable measurements are required. IB-OSC will be performed on the Thorasys TremoFLO device at each tertiary site. Participants will perform 3 x 20-second recordings of tidal breathing using a 10Hz sinusoidal waveform. All measurements will be performed by trained personnel, taking approximately 30-45mins.
iv. Mobility: The Gross Motor Function Measure (GMFM) item set version (GMFM- 66-IS) will be administered alongside the GMFM-66 (included in the Run4Health pilot study) to help capture changes in gross motor function across the spectrum of GMFCS levels (GMFCS II-V).^27^ The item set version uses an algorithm to determine which set of GMFM items is most appropriate based on participant’s level of function.
v. Frame Running activity and participation goal attainment: Children will set three goals related to Frame Running attendance, involvement and physical performance on the Canadian Occupational Performance Measure (COPM).^51^ Goal-directed intervention is considered best practice in rehabilitation,^52^ and enhances motivation to engage in physical activity.^53^ Test-retest reliability is high (ICC 0.76-0.89) and the measure is responsive with MCID=2 points.^51^
vi. Functional mobility: The Pediatric Evaluation of Disability Inventory Computer Adaptive Test (PEDI-CAT) mobility domain is a parent-report, standardised, norm-referenced assessment of mobility performance that is valid, reliable (ICC=0.98), responsive in CP and correlated with gross motor capacity.^28^
vii. Health related quality of life and mood: The KIDSCREEN-52 questionnaire is a 52 item quality of life questionnaire that is used extensively in children and adolescents (8-18 years) and young people with CP.^54^ A self-report and parent-proxy version of the questionnaire is available, and both versions are valid and reliable (ICC 0.56-0.77).^54^ The KIDSCREEN-52 questionnaire was developed to assess how children perceive their psychological, physical and social wellbeing.^55^
viii. Child Health Utility 9 Dimensions (CHU-9D) is a generic instrument for children up to 17 years giving a single preference-based utility index for health states, making the data amenable for economic evaluations of interventions.^56^ The EQ- 5D-5L is a generic health status questionnaire that measures health across five levels of severity which is commonly used in health economic evaluations.^57^ The EQ-5D-5L will be completed instead of the CHU-9D for participants aged 18 and older.
ix. Health Resource Utilisation: Participants will complete a Health Resource Utilisation Questionnaire (HRUQ) at study entry (T1) and study exit (T3 for intervention group, T4 for waitlist group), which will provide important information about the pattern of health care utilisation of young people with CP. The HRUQ will be modified (in consultation with the Run4Health consumer group) to reduce participant burden by removing any redundant/irrelevant items. The HRUQ will replace the usual care diary, previously included in the Run4Health pilot study. To reduce missing data, recall bias, and to more accurately determine costs incurred for accessing healthcare services, Medicare Benefits Schedule (MBS) and Pharmaceutical Benefits Scheme (PBS) data will also be obtained from Services Australia (following ethical approval and with informed participant consent). To estimate the true cost of the Run4Health intervention, the following data will be captured:

i. Direct medical costs: costs associated with directly providing the intervention, including diagnostic tests and professional staff time to deliver the intervention.
ii. Direct non-medical costs: non-medical resources required to deliver the intervention, such as transport, parking, track entry fees, etc.
iii. Overhead costs: administrative and support services required to deliver the intervention, such as equipment, supplies, and maintenance. Classification systems and demographic characteristics will be assessed and collected as per the Run4Health Pilot RCT protocol:^3^

i. Gross Motor Function Classification System Expanded and Revised (GMFCS)^58^
ii. Manual Abilities Classification System (MACS)^59^
iii. Communication Function Classification System (CFCS)^60^
iv. Visual Function Classification System (VFCS)^61^
v. Eating and Drinking Ability Classification System (EDACS)^62^
vi. Participant’s Frame Running Sport Class (RR1/RR2/RR3 and/or T71/T72) or provisional classification
vii. Participant age, sex, dominant hand and socioeconomic status
viii. Comorbid diagnoses
ix. List of up to nine sports/PAs the participant attended in the last 12 months
x. Caregiver frequency of participation in structured and unstructured sports/PAs in the last four months.

Participants will also be screened for medical conditions that may be precautions to high intensity exercise, requiring attention or adaptation but not meeting exclusion criteria (e.g. known stable cardiovascular or respiratory condition etc.).^1^

### Participant Timeline

Run4Health CP expanded schedule of assessments and interventions are provided below in Table 1 and the CONSORT^63^ study flow diagram is provided in Figure 1.

**Figure 1:**
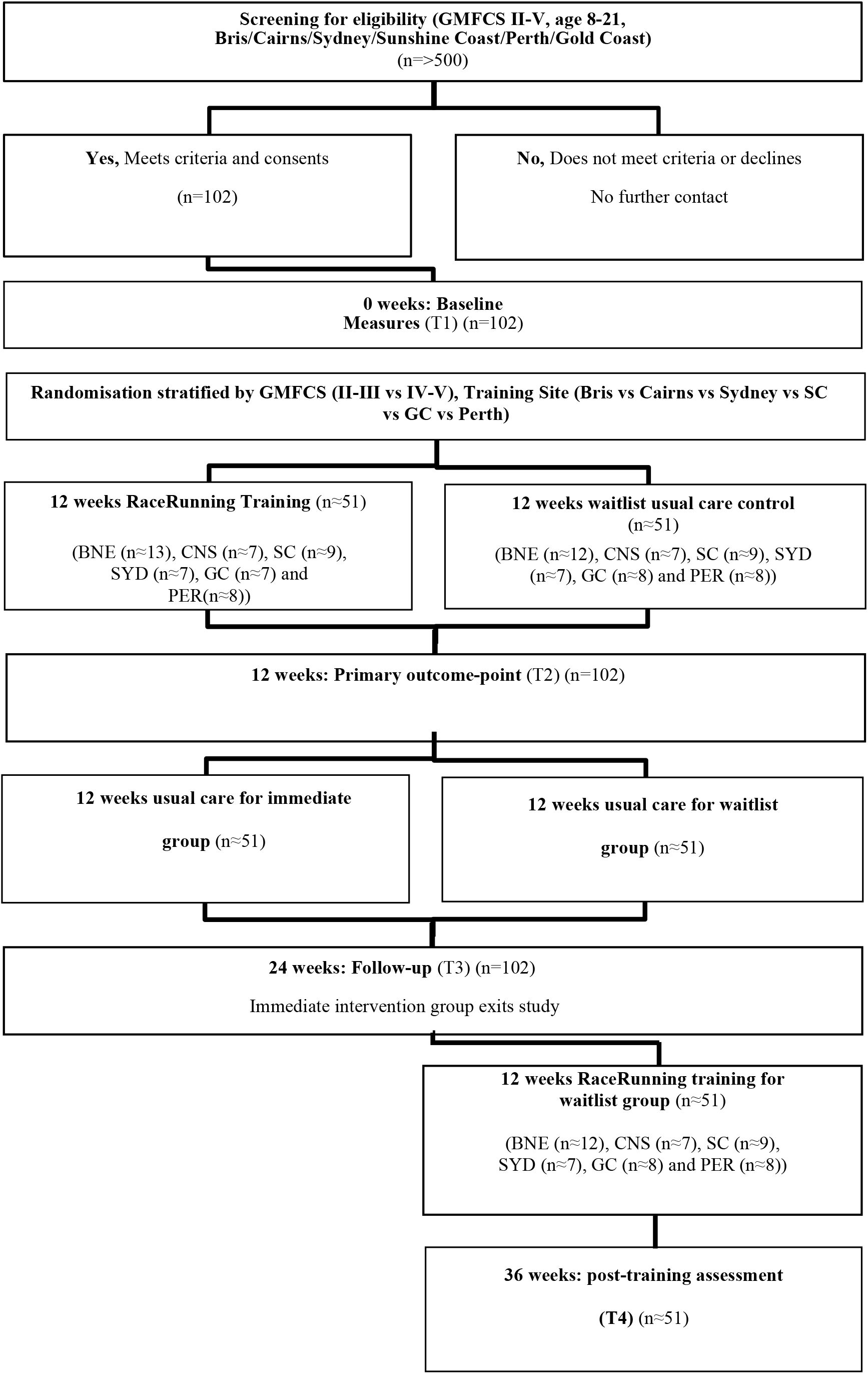
CONSORT Study Flow Diagram

**Table 1:** Updated schedule of assessments for Run4Health CP study.

| TIMEPOINT | Enrolment | Allocation/<br>Baseline | Intervention | Immediately<br>post-intervention<br>(12 weeks) | Retention (24<br>weeks) | Immediately<br>post-intervention<br>(waitlist group) |
| --- | --- | --- | --- | --- | --- | --- |
| VISIT NUMBER: | Screen | T1 |  | T2 | T3 | T4* |
| Participant contact | X |  |  |  |  |  |
| Participant screening (eligibility) | X |  |  |  |  |  |
| Informed consent | X |  |  |  |  |  |
| Medical & Physical Activity<br>questionnaire (Demographic<br>questionnaire) |  | X |  |  |  |  |
| Randomisation (group allocation) |  | X |  |  |  |  |
| INTERVENTIONS: |  |  |  |  |  |  |
| Frame Running training<br>(intervention) |  |  | X |  |  |  |
| Usual care (control) |  |  | X |  |  |  |
| **ASSESSMENTS:** |  |  |  |  |  |  |
| Participant classification form (GMFCS, MACS, CFCS, VFCS, EDACS, frame running class) |  | X |  |  |  |  |
| Frame Running provisional sport classification assessments (if required) |  | X |  |  |  |  |
| Gross Motor Function Measure 66 (GMFM-66) |  | X |  | X | X | X |
| Six-minute Frame Runner test (6MFRT) |  | X |  | X | X | X |
| Heart rate recovery in 1 minute (HRR<sub>1min</sub>) |  | X |  | X | X | X |
| Frame Running Functional Strength (activity limitation tests) |  | X |  | X | X | X |
| Resting blood pressure |  | X |  | X | X | X |
| Canadian Occupational Performance Measure (COPM) |  | X |  | X | X | X |
| Anthropometry (Body Mass Index – BMI, waist circumference) |  | X |  | X | X | X |
| Body Composition/DXA scan (% body fat/lean mass, bone mineral density [aBMD; g/cm<sup>2</sup>], bone mineral content [BMC; grams] at total body, AP lumbar spine, and lateral distal femurs) |  | X |  | X |  |  |
| Lung function tests: MIPS, MEPS, SNIPS, IB-OSC |  | X |  | X | X | X |
| Pediatric Evaluation of Disability Inventory Computer Adaptive Test (PEDI-CAT) mobility domain |  | X |  | X | X | X |
| Participation and Environment Measure for Children and Youth (PEM-CY) |  | X |  | X | X | X |
| Pittsburgh Sleep Quality Index (PSQI) |  | X |  | X | X | X |
| Child Health Utility 9 Dimensions (CHU-9D; participants <18 years) |  | X |  | X | X | X |
| EQ-5D-5L (participants >18 years) |  | X |  | X | X | X |
| KIDSCREEN-52 |  | X |  | X | X | X |
| 7-day free-living accelerometry for habitual physical activity and sleep data (ActiGraphGT3X+ thigh and wrist) |  | X |  | X | X | X |
| Wong-Baker FACES rating |  | X | X | X | X | X |
| Fatigue severity scale (FSS) |  | X | X | X | X | X |
| Visual Analogue Fatigue Scale (VAFS) |  | X |  | X | X | X |
| Health Resource Utilisation Questionnaire (HRUQ) |  | X |  |  | X** | X** |
| Adverse event monitoring and reporting |  | X | X | X | X | X |
| Participant interview at study exit |  |  |  |  | X** | X** |
| * T4 assessments will be completed by the waitlist control group only. The intervention group will exit the study following T3.<br>** Assessments will be completed at study exit. i.e. T3 for the intervention group and T4 for the waitlist control group. |  |  |  |  |  |  |

### Sample Size

The primary basis for sample size calculation is adequate power for H1 (6MFRT) comparison between functional effects of Run4Health CP compared to usual care immediately post intervention (T2). To be able to identify a between-group difference of 0.7 standard deviations or greater with 80% power (alpha=0.025 as 2-primary outcomes) at T2 require 40 participants in each group. This is equivalent to a MCID >148m for 6MFRT (assuming SD=220) given inclusion of GMFCS II-V and >13 for HRR1min (assuming SD=20). Based on previous RCTs conducted by our group we anticipate a maximum drop-out of 10%, we aim to recruit 90 children in total (45 in each group). To 17/05/2023, n=12 participants were enrolled in a pilot version of the trial. Data from these participants will be included in the analysis of the larger trial, allowing for a total of n=102 participants.

A minimum of 12 participants will be interviewed for the qualitative part of the study, with more recruited as time allows until there is saturation. Participants at the Brisbane site (minimum of 24 participants) will also be invited to participate in a Frame Running functional strength test reliability study, which will involve repeating the activity limitation tests two weeks apart to assess test-retest reliability. A minimum of 24 participants is below the recommended number of participants for a reliability study according to CONSORT guidelines however, given moderate-severe CP is a low-incidence disability and the logistical constraints, this will be a recognized limitation of the study.^3^

### Recruitment

Participants will be recruited across study sites as per the strategies outlined in the Run4Health Pilot protocol.^3^ Study flyers and information will be disseminated via clinical databases, clinical services (e.g. Queensland Paediatric Rehabilitation Service [QPRS] and Sydney Children’s Hospitals Network [SCHN]), patient waiting areas of the participating research sites, via clinical service newsletters, research websites, social media and word of mouth.

### Allocation and Blinding (Masking)

Participants will be randomly assigned to either the intervention group (Frame Running training) or waitlist control group (usual care) with a 1:1 allocation as per a computer-generated randomisation schedule using the Research Electronic Data Capture (REDCap®) randomisation module, stratified by GMFCS (II-III/IV-V) and training site (Brisbane vs Sunshine Coast vs Gold Coast vs Cairns vs Sydney vs Perth), using permuted blocks of random sizes. Randomisation will occur following enrolment into the study and completion of all baseline assessments except for 7-day habitual PA monitoring. Information about concealment and blinding (masking), who these apply to, how and when are provided in the Run4Health pilot RCT protocol.^3^ Procedures for emergency unblinding are not required, as participant health and safety is managed directly by Frame Running coaches who are not blind to treatment allocation.

### Data Collection

#### i) Interventionist Training and Experience

Frame Running sessions will be led by physiotherapists, exercise physiologists and/or athletic coaches with minimum experience of 2 years working with people with disabilities and delivering exercise programs to children and young people. Frame Running coaches will have current first aid and cardiopulmonary resuscitation (CPR) certifications and will adhere to institutional policies and procedures for child safety.^3^

Frame Running coaches will be provided with 1:1 training covering the following topics: i) general principles of aerobic and anaerobic exercise in CP, ii) coaching principles to provide a fun and intrinsically motivating exercise experience, iii) interpreting and applying the

Frame Running intervention manual, iv) correctly fitting athletes to running frames, and v) practical component. Regular supervision meetings will be conducted throughout the trial to facilitate adherence to the training manual.

#### ii) Assessor Training and Experience

Outcome assessors will be physiotherapists with at least 3 years’ experience working with children and youth with CP and will have completed the GMFM Criterion Test for scoring reliability. They will be provided with written and videotaped standardised procedures for the administration of all other study outcome measures. Regular supervision meetings will be conducted to facilitate adherence to the assessment manual.

### Retention

#### i) Participant Retention

Participant retention will be encouraged by: i) providing Frame Running training at no cost to the participant, ii) where possible, scheduling sessions at mutually convenient times, iii) administering questionnaires via the REDCap® survey module, iv) usual care control participants will be offered the intervention after the waitlist control period is complete (T3), v) once enrolled, investigators will encourage completion of overdue assessments by contacting participants and/or caregivers via their preferred method of contact (e.g. phone call, email, text messages).^3^

#### ii) Participant Withdrawal

Participants are free to withdraw from the study at any time without consequence, as outlined in the participant information sheet at study entry. Participants who choose to withdraw will be assisted to source another local therapy option that aligns with their preferences and goals, if desired. Deidentified data collected from participants who later withdraw (including re-identifiable data) will be retained and included in analyses. Reasons for participant withdrawal (if known) will be documented and reported.

### Data Management and Access

Patient-reported data (e.g. demographics, electronic questionnaires, etc) will be collected and managed directly via REDCap® (Research Electronic Data Capture) electronic data capture tools hosted at The University of Queensland.^64 65^ Paper based data (objective measures such as the GMFM) will be completed in-person by study assessors and later digitized and transferred to the University of Queensland Research Data Manager (UQRDM) database for long term data storage. Raw scores from objective assessments will be recorded in REDCap®. Habitual physical activity and heart rate data will be recorded using wearable devices and the deidentified data will be transferred to the UQRDM database (via the required proprietary software) for storage. Data will be erased from the wearable devices once uploaded to the UQRDM. Data from interviews recorded via ZOOM web conferencing software or video camera will be saved electronically on the UQRDM database as soon as practical following the interview, and then erased from the recording device. Most data will be collected in a de-identified (but re-identifiable) format, with the exception of photographs, videos, and voice recordings which are identifiable. Confidentiality of participant data will be maintained at all times from collection to storage. A de-identified dataset will be made available upon written request for the purposes of further scientific research, including meta-analysis, ancillary studies related to the original aims and objectives, and verification of results.

### Statistical Methods

Analyses will follow standard principles for RCTs using two-group comparisons using parametric statistics on all participants on an intention-to-treat basis. Primary comparison immediately post intervention (T2) will be based on distance on 6MFRT. Groups will be compared using a linear regression model, with study group (intervention/control) included as the main effect. The effect estimate will be presented as mean difference and 95% confidence interval with a significance level of p<0.05. Secondary outcomes with interval data will be examined using linear regression models, with binary data using logistic regression models and with count data using Poisson regression models. If model assumptions are not met non-parametric alternative models will be used. When repeated measures data is analysed, mixed-effects models with participant included as a random effect will be used as appropriate.

Training data from the intervention and waitlist group will be pooled in a secondary analysis at three timepoints (baseline, immediately post-training and 12 weeks post-training) to investigate pre-post intervention changes.

Qualitative interview transcripts will be thematically analysed (using Theoretical Domains Theory as a guiding framework)^29^ following transcription with a content analysis approach using NVivo. The test-retest, intra-rater and interrater reliability study for Frame Running functional activity limitation tests will be analysed using accepted parametric methods where appropriate (i.e. Standard Error of Measurement and Intraclass Correlation Coefficients). Construct validity of the Frame Running functional activity limitation tests will also be assessed using Pearson’s r correlation coefficient to examine convergent validity with 6MFRT and known-groups validity using Frame Running classification and GMFCS level.

### Data Monitoring and Safety

By their nature, sports and active recreation activities may have small to moderate risks of injury associated with participation due to hazards present (some of which are integral parts of the activity and cannot be removed). There are also negligible to small risks of psychological harm associated with disclosure of personal/sensitive information. Furthermore, during the study therapists will use common, approved devices in the course of their work for approved purposes, including ActiGraph accelerometers, Axivity AX3 monitors and running frames, which may have some inherent risks associated. To minimize risk of injury and adverse events, the following control measures will be implemented: (1) participant screening for comorbid medical conditions and risk factors, (2) information sheets and counselling will be provided to participants and families on the risks associated with wearing accelerometers (pressure areas, allergic skin reactions, etc), (3) standardised training will be provided to study assessors and coaches, (4) participants must wear appropriate footwear and use a properly fitted bicycle helmet (compliant with Australian standards), (5) participants will have access to clean drinking water and be encouraged to use sun protection during training sessions, (6) a familiarization session will be provided to all participants, to instruct them on the safe use of the running frame (at least 10 minutes), and (7) pain and fatigue will be continually monitored and the training load will be adapted accordingly. Adverse events will be reported by coaches and assessors via REDCap® as soon as practical following an incident or adverse report. Adverse events will be monitored by the principal investigator, and serious or unexpected events will be discussed at the earliest opportunity by the chief investigators (SR, LS, SO, MA, EA, AB, TB, SK, ID, DP) and reported to the ethics committee. An independent Data Safety Monitoring Committee (DSMC) comprising leading clinicians, trial experts and statistician will be established to ensure patient safety and trial and data integrity. The DSMC will meet twice per year and be on call to assess serious adverse events.

### Qualitative Interviews

At study exit (T3 for the intervention group, T4 for the waitlist group), participants and caregivers will be invited to participate in a qualitative interview. The purpose of the interview is for participants to share their experiences in the training program, including any perceived facilitators or barriers to participation (and ongoing participation) in Frame Running. Interviews will be completed and analysed as per methods outlined in the Run4Health pilot RCT protocol.^1^

### Consumer and Public Involvement

A Run4Health consumer group, consisting of athletes with cerebral palsy and their caregivers will be established to provide ongoing guidance and input throughout the research process. Consumer representatives will be involved at all research stages, including co-development and review of the trial protocol, participant information and consent materials, assessment materials and procedures, implementation, and dissemination of results. Run4Health consumer representatives will be financially compensated for their time and expertise. Consumer representatives will nominate their preferred method of payment (either direct credit into their account or gift card). The rate of reimbursement will be equivalent to the Health Consumer Queensland (HCQ) remuneration rates^66^, adjusted for annual inflation increases (1.5%). Consumer council meetings will be held no less than twice yearly, and one consumer representative will be invited to sit on the trial management committee.

## ETHICS AND DISSEMINATION

### Informed Consent Process

Written informed consent will be obtained from legal guardians for children under the age of 18 years, and for participants who are older than 18 years with impaired capacity to consent. Participants who are older than 18 years and have capacity to provide their own written informed consent will be asked to do. Consent will be gained after the participant and caregiver have received the written study information sheet and the treating/assessing staff member has explained the study to the satisfaction of the participant and legal guardian.

### Ethics and Dissemination

The expanded Run4Health CP RCT has been approved by the Children’s Health Queensland Hospital and Health Service Human Research Ethics Committee (HREC/21/QCHQ/69281) and the University of Queensland Human Research Ethics Committee (2021/HE000725). The trial is also registered on the Australian New Zealand Clinical Trials Registry (ANZCTR, ACTRN12621000317897). The trial registration will be amended to reflect any protocol updates, and deviations from the protocol will be reported in the primary results manuscript. Study outcomes will be disseminated via the ANZCTR website, conference presentations and abstracts, peer-reviewed scientific journals, organization/institution media releases and newsletters and directly to research participants in an appropriate and accessible format.

## AUTHORS’ CONTRIBUTIONS

The original pilot RCT was conceived by SR, LS, LM, CS, EB, KW and RNB. The expanded RCT protocol, funded by a MRFF early career research grant, was conceived by SR, LS, SO, MA, EA, AB, TB, SAK, ID, DP & RNB. EA & SR completed the initial draft of the manuscript. LGP co-ordinated the pilot study and made significant contributions to the expanded study protocol. RW provided biostatistical advice and information. SGT, MA, RT, and ID contributed technical expertise to the protocol manuscript for physical activity and sleep measurement, therapist outcome assessment, and Frame Running coaching respectively. SO and DB contributed technical expertise to the protocol manuscript and assessment manual for DXA procedures and assessment, and TB provided technical expertise regarding respiratory function assessments. SAK and TC provided expert guidance on health economic data analysis methods. All authors designed the study, have read, edited, and approved the final manuscript and supplementary files.

## FUNDING STATEMENT

The Run4HealthCP trial is financially supported by an Early Career Research Project Support Grant awarded by the Children’s Hospital Foundation (grant ID: ECR0262020), the Dr June Canavan Foundation, the Merchant Charitable Foundation via the Children’s Hospital Foundation (donation ID: 10415), the Medical Research Future Fund (MRFF) via National the Australian National Health and Medical Research Council (NHMRC) 2021 Early to Mid-Career Researchers grant (2022624) and NHMRC Research Fellowship grant (RNB).

Assistive technology supplier Dejay Medical Pty Ltd has agreed to provide at least a 30% discount on Running Frames required to be purchased for the trial. Other running frames used in the trial will be borrowed at no cost (except return freight) from various individuals and organisations.

## Data Availability

Data produced from the present study will be disseminated in scientific journals (manuscripts) on study completion.

## Footnotes

1 Run4Health Doseand risk assessment report v2

